# Long COVID Prevalence among U.S. Adults: A State-level Ecological Analysis of the Contribution of COVID-19 Incidence, Severity of Acute Illness, COVID-19 Vaccination, and Chronic Conditions

**DOI:** 10.64898/2026.03.07.26347841

**Authors:** Xinmeng Zhao, Li Deng, Nicole D. Ford, Sharon Saydah

## Abstract

**Background:** Long COVID has emerged as a major public-health concern in the United States, yet geographic variation in its prevalence remains poorly understood. This study examines how state-level differences in COVID-19 vaccination, SARS-CoV-2 incidence, COVID-19 hospitalization, and chronic disease burden relate to adult Long COVID prevalence in the United States.

**Methods:** We conducted an ecological analysis using data from the 2023 Behavioral Risk Factor Surveillance System (BRFSS), from which we estimated state-level prevalence of self-reported Long COVID among adults. These estimates were linked with publicly available data on SARS-CoV-2 incidence, COVID-19 hospitalizations, COVID-19 vaccine coverage, and a multimorbidity indicator (>= 3 chronic conditions e.g., diabetes, obesity, chronic kidney disease) associated with higher risk for severe SARS-CoV-2. Multivariable linear regression models were fitted to assess the contribution of each factor adjusted for age and sex distribution, incorporating Rubin’s rules to account for uncertainty in prevalence estimates.

**Results:** All examined factors—including SARS-CoV-2 incidence, hospitalization rates, and multimorbidity, vaccine coverage—varied by state. When modeled simultaneously and adjusting for age and sex distribution, only COVID-19 vaccine coverage and SARS-CoV-2 incidence were significantly associated with Long COVID prevalence. COVID-19 vaccine coverage showed a strong protective association, while SARS-CoV-2 incidence showed a modest positive association. Multimorbidity and hospitalization rates were not independently associated with adjustment.

**Conclusions:** State-level variation in Long COVID burden appears most strongly driven by COVID-19 vaccine coverage and SARS-CoV-2 incidence. Promoting COVID-19 vaccination remains essential to reduce long-term impacts from SARS-CoV-2 infections.

## Introduction

### Background

Long COVID, defined as new or persisting symptoms such as fatigue, headache, dyspnea and anosmia that last three or more months after acute SARS-CoV-2 infection [1], has emerged as a major public health concern in the United States. According to the 2023 Behavioral Risk Factor Surveillance System (BRFSS), age- and sex-standardized adult Long COVID prevalence ranged widely across the United States [2]. Similar levels of geographic heterogeneity have been documented across prior BRFSS cycles and other national surveys [3], highlighting consistent state-to-state variation in adult Long COVID prevalence.

Certain groups are more likely to experience Long COVID, particularly individuals who had more severe acute COVID-19 [4], were unvaccinated [4–6], or had pre-existing chronic conditions—such as diabetes, heart disease, and obesity [4,7,8]. These factors may contribute to the observed geographic variation in Long COVID prevalence. While many studies have examined individual-level risk factors for Long COVID, few have attempted to quantify associations with Long COVID prevalence at the population level. It remains unclear whether geographic variation in Long COVID prevalence reflects underlying differences in these factors. Understanding these associations is critical for helping state and local health departments identify and support populations at higher risk and develop targeted prevention strategies.

Currently, there is no integrated dataset that captures all relevant individual-level factors associated with Long COVID across all U.S. states [3]. Given these limitations, an ecological approach that leverages publicly available, aggregate data provides a policy-relevant alternative. By synthesizing state-level indicators of infection burden, acute illness severity, vaccine coverage, and comorbidity prevalence, it is possible to assess whether the observed geographic variation in Long COVID prevalence can be partially explained by underlying differences in these indicators. This approach is especially valuable for guiding public health interventions in the absence of extensive datasets with person-level data.

The objective of this study is to assess the association of state-level geographic variation in adult Long COVID prevalence with SARS-CoV-2 incidence, acute COVID-19 illness severity, COVID-19 vaccine coverage, and prevalence of chronic conditions.

## Methods

### Study Design and Setting

We conducted an ecological study of adult Long COVID prevalence across U.S. states, using the state as the geographic unit. The primary aim was to identify factors associated with geographic variation in Long COVID burden using publicly available, population-level data sources.

### Outcome measure

The primary outcome was the crude, state-level prevalence of Long COVID among adults, based on data from 2023 Behavioral Risk Factor Surveillance System (BRFSS) [9], with data collection extending into early 2024 (January through March, depending on the state). BRFSS is a state-based system of telephone health surveys that collects information on health risk behaviors, preventive health practices, and health care access. It is designed to produce representative estimates for non-institutionalized adults aged 18 years and older in participating states and territories. Respondents were asked if they ever tested positive for COVID-19 or had been told by a doctor or other health care provider that they have or had COVID-19. Those who provided an affirmative response were then asked if they experienced symptoms lasting 3 months or longer that were not present prior to having COVID-19. If yes, respondents reported whether they were still experiencing symptoms at the time of interview (i.e., current Long COVID).

Final BRFSS sampling weights were applied to compute state-specific prevalence estimates and standard errors to make the sample statistics representative of the state or territory population from which it was drawn. The 2023 BRFSS public-use dataset excluded Kentucky and Pennsylvania due to insufficient data collection, resulting in a final analytic dataset comprising 48 states.

### Potential State-Level Factors Associated with Long COVID

To align with the 2023 BRFSS survey timing and the definition of Long COVID, we defined the analytic window as October 1, 2022, to September 30, 2023. This interval was selected to capture factors potentially associated with Long COVID preceding and overlapping with the timeframe relevant to the Long COVID definition, ensuring temporal alignment with prevalence measured in 2023. Final variables were selected based on data availability and represent the best feasible synthesis across the full analytic period and geographic scope. A summary of each data source is provided in Supplementary Table 1, with an overview presented below.

To quantify SARS-CoV-2 incidence, we obtained state-level counts of new COVID-19 cases (confirmed and probable cases) from the Centers for Disease Control and Prevention (CDC) COVID-19 Response Data [10]. Mandatory COVID-19 reporting requirements ceased after May 10, 2023, following the official declaration of the end of the public health emergency. Iowa stopped reporting earlier, on April 1, 2023, and five additional states missed one week of reporting. To account for this gap, we aggregated incident cases between October 1, 2022, and May 10, 2023, and calculated incidence rates as cumulative new cases per person-year, using 2022 state population estimates [11].

To capture acute COVID-19 illness severity, we used state-specific weekly adult COVID-19 hospitalization rates per 100,000 persons computed based on data from the National Healthcare Safety Network (NHSN) [12]. NHSN is a national surveillance system that collects real-time hospitalization data from participating facilities across the United States. There was no missing data for hospitalizations. For comparability, we calculated the cumulative hospitalization rate over the full analytic period (October 1, 2022, to September 30, 2023) and converted it to cumulative new hospitalizations per person-years.

For COVID-19 vaccine coverage we used monthly, state-specific estimates from the National Immunization Survey (NIS) [13], a set of telephone-based surveys designed to monitor vaccination coverage in children, adolescents, and adults in the United States. Specifically, we extracted data from the NIS–Adult COVID Module (NIS-ACM), which reports the proportion of adults who received a COVID-19 bivalent booster vaccine following completion of the primary vaccine series. This measure captures recent vaccination during the analytic period rather than historical vaccine coverage (i.e., ever vaccinated). Emerging evidence suggests that more recent COVID-19 vaccination is associated with a reduced risk of developing Long COVID [14]. Cumulative COVID-19 vaccine coverage was calculated as the weighted average of monthly percent booster coverage among survey respondents who had completed the primary vaccine series. The weights were based on the number of respondents with completed primary series in each month between October 2022 and June 2023.

For chronic conditions, we used a composite comorbidity indicator representing the proportion of adults with three or more chronic health conditions, hereafter referred as “multimorbidity”, from unpublished 2024 estimates produced by the Center for Disease Control and Prevention’s Center for Forecasting and Outbreak Analytics (CFA). The estimates synthesized data from multiple data sources including 2022-2023 medical claims, consumer records, BRFSS, and the National Health Interview Survey (NHIS) data to better estimate state-level comorbidity prevalence and covered a broader set of conditions relevant to severe COVID-19. The comorbidity conditions included asthma, obesity, diabetes, heart condition, current or former smoking, tuberculosis, cerebrovascular disease, chronic liver disease, chronic kidney disease, chronic lung disease, mental health disorder, cystic fibrosis, HIV, immunodeficiency, transplantation, immunosuppression, cancer, pregnancy, disability and neurocognitive dementia. Detailed methods are available upon request.

Although BRFSS allows for age- and sex-standardized estimation of Long COVID prevalence—and similar standardization was possible for multimorbidity—comparable adjusted data were unavailable for other factors (e.g., state-level COVID-19 cases and hospitalization rates). Consequently, all factors and the outcome variable were retained in their crude (unstandardized) form, and each state’s population structure was captured with three variables derived from the U.S. Census’ 2022 American Community Survey [11]: (i) the proportion of adults aged ≥ 65 years, (ii) the proportion of male adults aged >= 65 years, and (iii) the proportion of female adults aged < 65 years. Together, these variables reflect both the independent distributions of age and sex as well as conditional age–sex subgroups.

### Statistical Analysis

We computed descriptive statistics for Long COVID prevalence and all factors, including cumulative incidence per person-years, cumulative hospitalizations per person-years, vaccine coverage, multimorbidity). We then conducted univariable linear regressions to assess the association between Long COVID prevalence and each factor individually. Finally, we fit a multivariable model that incorporated all four state-level factors along with the three age and sex variables defined above. We presented regression coefficient (β) and its 95% confidence interval (CI), where β is interpreted as the average magnitude of change in Long COVID prevalence with one unit increase in a given factor.

State-level Long COVID prevalence was derived from survey estimates with known standard errors, so we propagated that sampling uncertainty (i.e., survey error) through all subsequent modeling. Specifically, we generated 1,000 Monte-Carlo draws from a normal distribution with BRFSS mean prevalence estimate and the corresponding standard error. Each draw yields a complete prevalence vector for the 48 states and thus a new analytic data set. We fit the full multivariable model to each data set, retained the regression coefficients, and then combined estimates using Rubin’s rules [15], thereby producing pooled point estimates and total variances that incorporated both within- and between-simulation variability. COVID-19 vaccine coverage was also sourced from survey-based data (NIS–ACM); however, this survey dataset provided uncertainty to monthly/weekly (check) responses only. No cumulative vaccine coverage uncertainty was available for this analysis. So, we did not add the uncertainty of the vaccine coverage estimate in this analysis. No information on uncertainty was available for the other factors (e.g., SARS-CoV-2 incidence, COVID-19 hospitalizations and multimorbidity) considered.

Prevalence of Long COVID was estimated from BRFSS survey, using SAS® software, version 9.4 (SAS Institute Inc., Cary, NC). Statistical modeling and simulation-based inference were conducted in R version 4.4.0 (R Foundation for Statistical Computing, Vienna, Austria). Statistical significance was defined as p < 0.05, with results at p < 0.01 and p < 0.001 noted as stronger levels of significance.

### Ethical considerations

This study used de-identified, publicly available secondary data. This activity was reviewed by CDC, deemed not research, and conducted consistent with applicable federal law and CDC policy (45 C.F.R. part 46.102(l)(2), 21 C.F.R. part 56; 42 U.S.C. Sect. 241(d); 5 U.S.C. Sect. 552a; 44 U.S.C. Sect. 3501 et seq.).

## Results

### Descriptive Statistics

During the analytic window of this analysis (October 2022–Sep 2023; Figure 1), prevalence of Long COVID displayed interstate heterogeneity, ranging from 4.8% in Maryland to 9.7% in West Virginia, with a median of 6.6% (Q1–Q3 5.9%–7.3%) (Figure 2). State-level factors likewise varied across the 48 states (Table 1). Cumulative SARS-CoV-2 incidence per person-years averaged 0.05 (SD = 0.01), with a median of 0.05 (Q1–Q3: 0.043–0.058) and a range of 0.034–0.076. Cumulative COVID-19 hospitalizations per person-years were lower in magnitude but similarly heterogeneous, averaging 0.003 (SD = 0.001), with an interquartile range tightly centered around 0.003 and spanning 0.002–0.004. COVID-19 vaccine coverage was more variable, averaging 28.9% (SD = 5.2%), with a median of 28.9% (Q1–Q3: 25.0%–32.7%) and a range of 18.8%–39.3%. Finally, multimorbidity (at least 3 comorbidity conditions) averaged 18.6% (SD = 3.9%), with a median of 18.8% (Q1–Q3: 15.8%–21.3%) and a range of 10.8%–29.0%.

**Figure 1.**
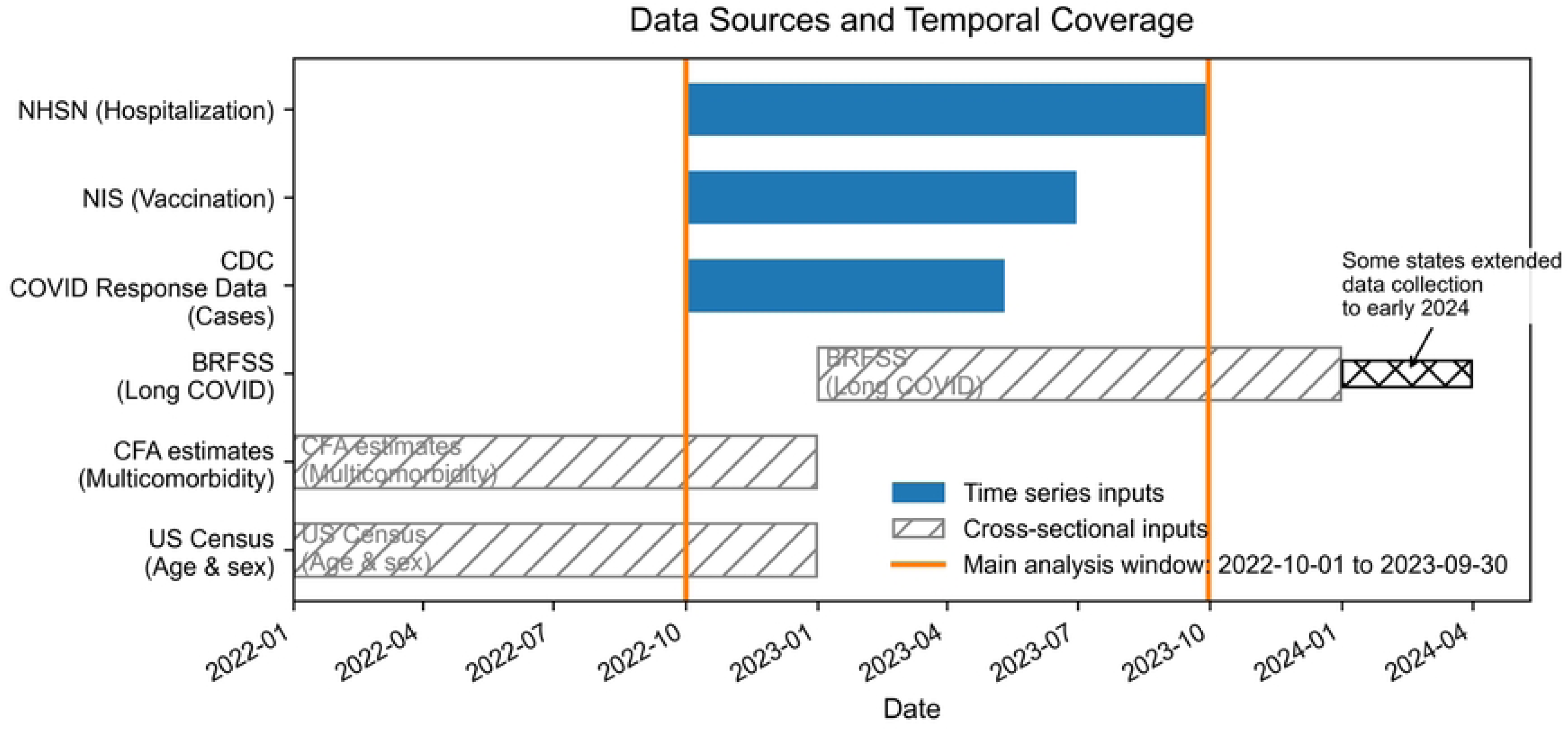
Temporal Coverage of Data Sources and Main Analysis Window. Solid bars represent time series inputs (e.g., hospitalizations, vaccination, cases), while hatched bars indicate cross-sectional data sources (e.g., demographics, multimorbidity, long COVID estimates). The orange vertical lines denote the start and end of the main analysis period (October 2022 to October 2023).

**Figure 2.**
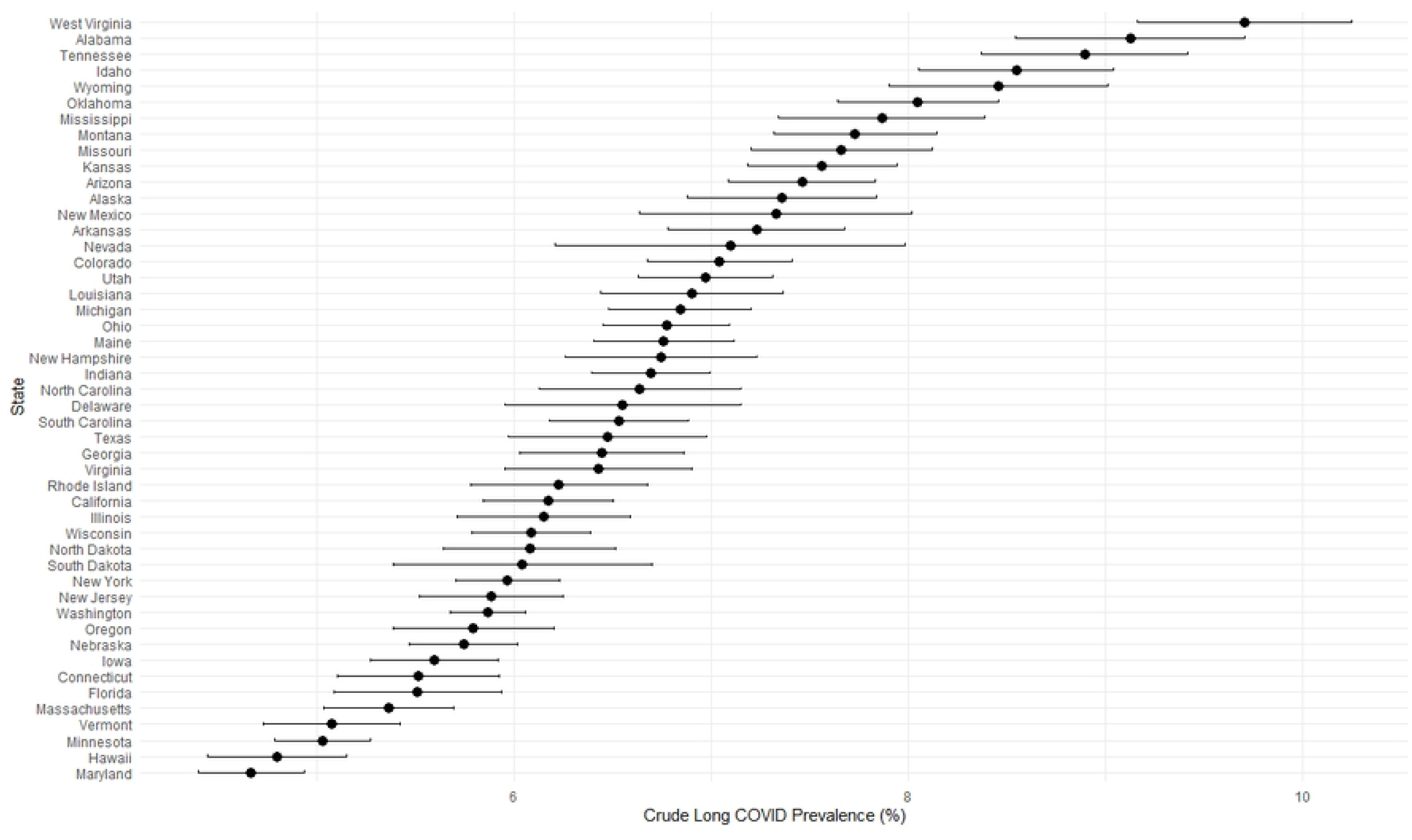
Prevalence of Long COVID among Adults by State with 95% CI. Estimated prevalence of Long COVID among U.S. adults by state in the 2023 BRFSS [9], ordered from highest to lowest prevalence. Error bars represent 95% CI of the estimates derived from the 2023 BRFSS data. Kentucky and Pennsylvania were not included in the public-use 2023 BRFSS data set. Non-state jurisdictions were also excluded from analysis. Abbreviations: BRFSS, Behavioral Risk Factor Surveillance System; CI, confidence interval.

**Table 1.** Descriptive Statistics of Long COVID Prevalence and State-Level Factors State-Level.

| State-Level Factor | Mean (SD) | Median (Q1-Q3) | Range |
| --- | --- | --- | --- |
| Long COVID prevalence | 0.067 (0.011) | 0.066<br>[0.059 – 0.073] | 0.047 – 0.097 |
| SARS-CoV-2 incidence per person-years | 0.05 (0.01) | 0.05<br>[0.043 – 0.058] | 0.034 – 0.076 |
| COVID-19 hospitalizations per person-years | 0.003 (0.001) | 0.003<br>[0.003 – 0.003] | 0.002 – 0.004 |
| COVID-19 vaccine coverage | 0.289 (0.052) | 0.289<br>[0.25 – 0.327] | 0.188 – 0.393 |
| ≥3 chronic conditions | 0.186 (0.039) | 0.188<br>[0.158 – 0.213] | 0.108 – 0.29 |

Data were obtained from CDC COVID-19 Response Data (SARS-CoV-2 incidence) [10], NHSN (COVID-19 hospitalization rates) [12], NIS-ACM (COVID-19 vaccine coverage) [13], and CDC CFA (≥3 chronic conditions).
Abbreviations: BRFSS, Behavioral Risk Factor Surveillance System; NHSN, National Healthcare Safety Network; NIS-ACM, NIS–Adult COVID Module; CDC, Centers for Disease Control and Prevention; CI, confidence interval.

### Univariable Regression

In univariable regressions (Table 2), vaccine coverage and multimorbidity were significantly associated with Long COVID prevalence. States with higher vaccine coverage tended to have lower Long COVID prevalence (β = −0.13; p < 0.001). Similarly, states with a higher proportion of adults with 3 or more chronic conditions observed higher Long COVID prevalence (β = 0.13; p = 0.001). Cumulative SARS-CoV-2 incidence per person-years and hospitalization per person-years were not significantly related to Long COVID prevalence (both p > 0.05). Figure 3 presents the scatterplot of vaccine coverage against Long COVID prevalence, suggesting a negative linear association between Long COVID prevalence and booster coverage at the state level (i.e., states with higher COVID-19 vaccine coverage tend to have lower Long COVID prevalence).

**Figure 3.**
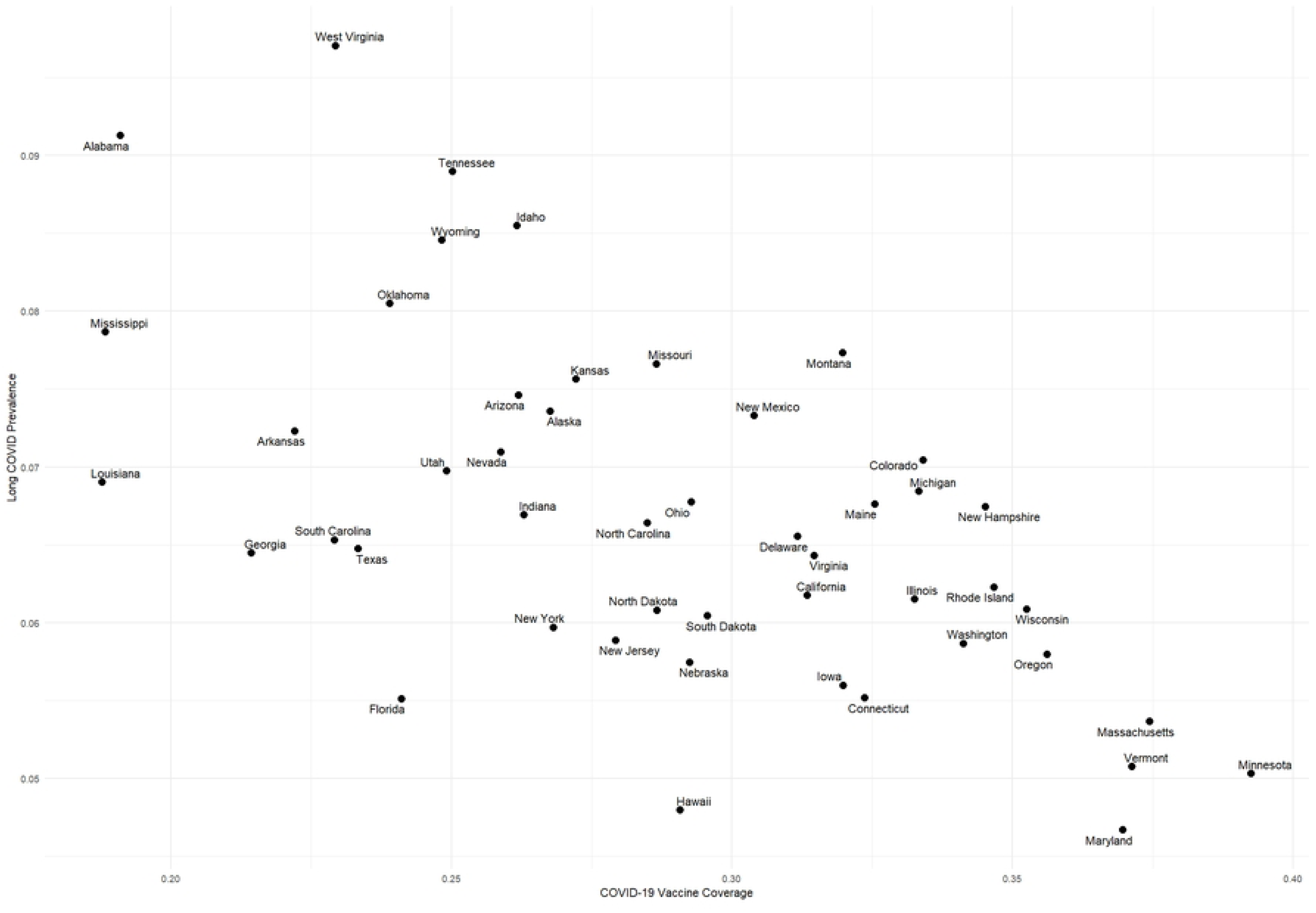
Scatterplot of Adult Long COVID Prevalence by Cumulative Vaccine Coverage by State, 2023. Long COVID data are from the 2023 BRFSS [9]. Kentucky and Pennsylvania were not included in the BRFSS public-use dataset; vaccine coverage data are from the NIS-ACM [13]. Abbreviations: BRFSS, Behavioral Risk Factor Surveillance System; NIS-ACM, NIS–Adult COVID Module.

**Table 2.**
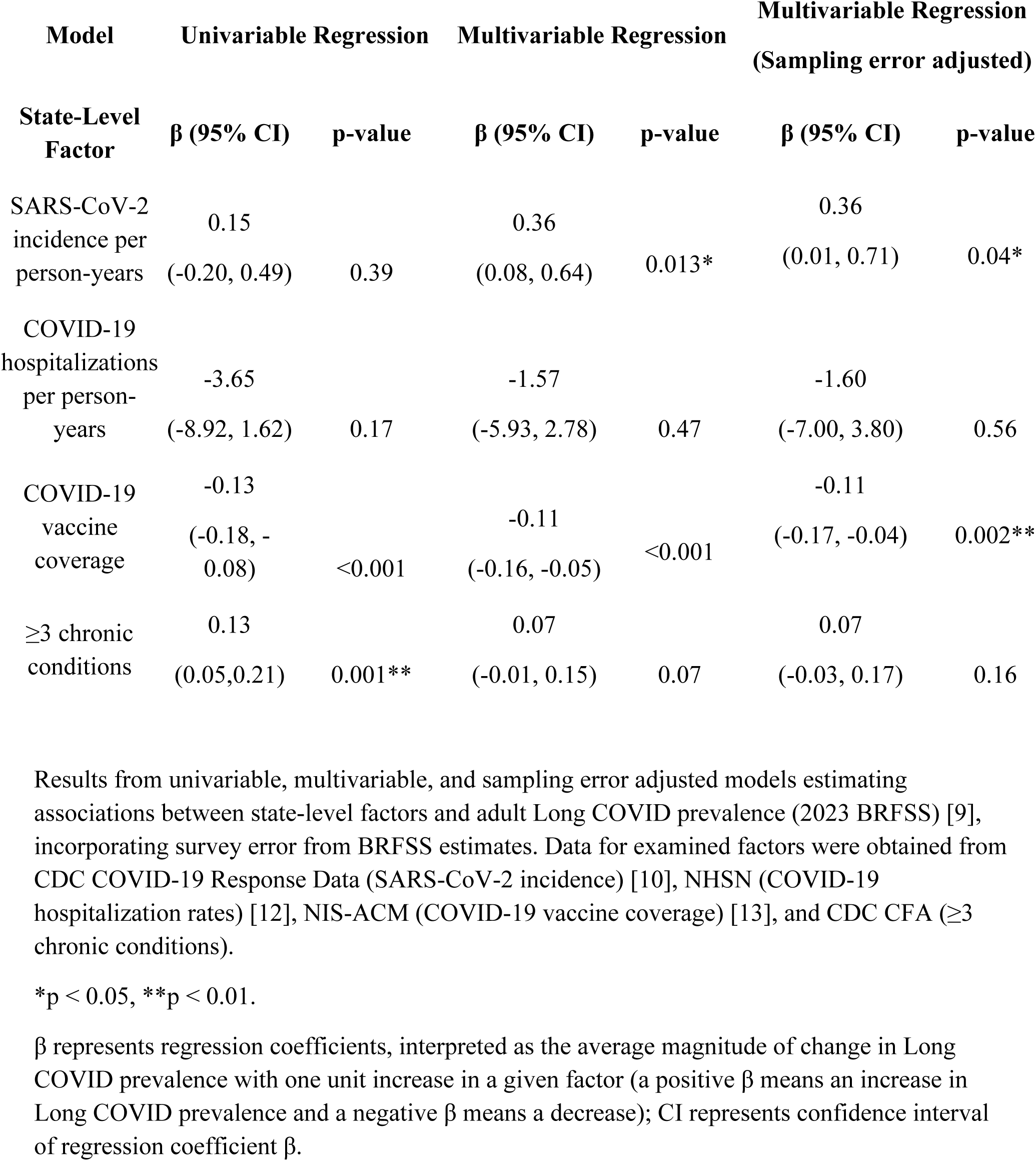
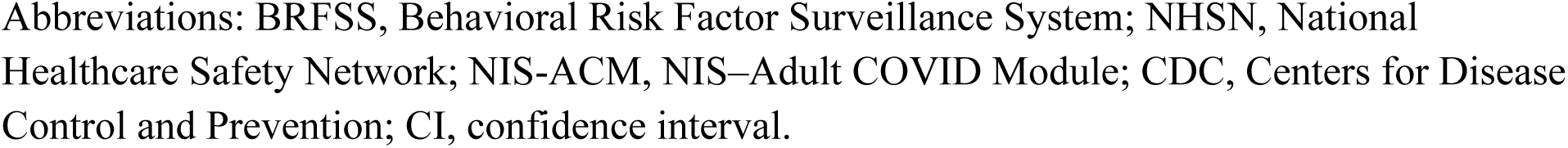
Associations of State-Level Factors with Long COVID Prevalence Across Univariable, Multivariable, and Sampling–Error–Adjusted Models of Adult Long COVID Prevalence.

| Model | Univariable Regression |  | Multivariable Regression |  | Multivariable Regression<br>(Sampling error adjusted) |  |
| --- | --- | --- | --- | --- | --- | --- |
| State-Level Factor | $\beta$ (95% CI) | p-value | $\beta$ (95% CI) | p-value | $\beta$ (95% CI) | p-value |
| SARS-CoV-2 incidence per person-years | 0.15<br>(-0.20, 0.49) | 0.39 | 0.36<br>(0.08, 0.64) | 0.013* | 0.36<br>(0.01, 0.71) | 0.04* |
| COVID-19 hospitalizations per person-years | -3.65<br>(-8.92, 1.62) | 0.17 | -1.57<br>(-5.93, 2.78) | 0.47 | -1.60<br>(-7.00, 3.80) | 0.56 |
| COVID-19 vaccine coverage | -0.13<br>(-0.18, -0.08) | <0.001 | -0.11<br>(-0.16, -0.05) | <0.001 | -0.11<br>(-0.17, -0.04) | 0.002** |
| $\geq 3$ chronic conditions | 0.13<br>(0.05, 0.21) | 0.001** | 0.07<br>(-0.01, 0.15) | 0.07 | 0.07<br>(-0.03, 0.17) | 0.16 |

Results from univariable, multivariable, and sampling error adjusted models estimating associations between state-level factors and adult Long COVID prevalence (2023 BRFSS) [9], incorporating survey error from BRFSS estimates. Data for examined factors were obtained from CDC COVID-19 Response Data (SARS-CoV-2 incidence) [10], NHSN (COVID-19 hospitalization rates) [12], NIS-ACM (COVID-19 vaccine coverage) [13], and CDC CFA ( $\geq 3$ chronic conditions).
\* $p < 0.05$ , \*\* $p < 0.01$ .
$\beta$ represents regression coefficients, interpreted as the average magnitude of change in Long COVID prevalence with one unit increase in a given factor (a positive $\beta$ means an increase in Long COVID prevalence and a negative $\beta$ means a decrease); CI represents confidence interval of regression coefficient $\beta$ .
Abbreviations: BRFSS, Behavioral Risk Factor Surveillance System; NHSN, National Healthcare Safety Network; NIS-ACM, NIS–Adult COVID Module; CDC, Centers for Disease Control and Prevention; CI, confidence interval.

### Multivariable Regression

In the multivariable model including all four state-level factors and adjusted for age and sex distribution (proportion ≥ 65 years old; proportion male ≥ 65 years; proportion female < 65 years), vaccine coverage remained negatively associated with Long COVID prevalence (β = – 0.11, p < 0.001) (Table 2). Cumulative SARS-CoV-2 incidence per person-years was positively associated with Long COVID prevalence after adjustment (β = 0.36; p = 0.013). Multimorbidity fell short of significance (β = 0.07, p = 0.07); this attenuation might be due to correlation with COVID-19 coverage. COVID-19-hospitalization rates were not associated with Long COVID prevalence with full adjustment.

### Uncertainty Analysis

The results using the Monte Carlo simulations were similar to those from the multivariable regression using point estimates of Long COVID prevalence without standard error incorporated in the outcome (Table 2); COVID-19 vaccine coverage remained significantly negatively associated with Long COVID prevalence (mean β = −0.11; p < 0.01) while SARS-CoV-2 incidence per person-years was weakly associated with Long COVID prevalence (mean β =0.58; p=0.038). The other state-level factors, multimorbidity and hospitalizations were not associated with Long COVID prevalence.

## Discussion

There are two major findings in this ecological study. First, in addition to Long COVID prevalence, all state-level factors we examined, vaccine coverage, SARS-CoV-2 incidence, COVID-19 hospitalizations, and multimorbidity, showed marked interstate variation. Second, when these factors were examined together in a multivariable model, only vaccine coverage and SARS-CoV-2 incidence were significantly associated with Long COVID prevalence, after adjusting for sex and age variables. Higher vaccine coverage was associated with a lower Long COVID prevalence, whereas greater cumulative SARS-CoV-2 incidence showed a modest association with Long COVID prevalence. These findings were robust to uncertainty analyses which accounted for margin of error in state-level Long COVID estimation. Our findings highlight the importance of COVID-19 vaccination and SARS-CoV-2 infection prevention in mitigating adult Long COVID. These results may inform state and local health departments in identifying and supporting populations at higher risk and support development of targeted prevention strategies.

A growing body of cohort studies and meta-analyses demonstrates that recent vaccination lowers the probability of developing Long COVID [5,14,16,17]. Our findings were consistent with the body of evidence. The negative correlation between vaccine coverage and Long COVID prevalence persisted after controlling for all other factors. Some studies have reported conflicting findings [18,19], which may be due to differences in Long COVID definitions, follow-up intervals, or dominant variant.

Individuals with multiple chronic conditions are more likely to have severe outcome after SARS-CoV-2 infection[20]. In our study, multimorbidity was positively associated with Long COVID prevalence in the univariable model but not after adjustment for all other state-level factors and age and sex variables. The attenuation of the relationship between multimorbidity and adult Long COVID prevalence at the state level may reflect shared pathways through severe acute COVID-19 disease risk, vaccine coverage or age and sex variables, which were already captured in our multivariable analysis.

At the individual-level, severe acute COVID-19 disease, including hospitalizations, has been shown to increase the risk of developing Long COVID [4,21]. Our study did not find significant association between cumulative COVID-19 hospitalization rates and Long COVID prevalence - with or without adjustment. There are several possibilities for this discrepancy. First, it may in part reflect the ecological nature of our analysis, as associations observed at the individual level may not translate directly to population-level patterns. Additional factors may also contribute. Because BRFSS includes only the non-institutionalized adult population, individuals discharged to long-term care or those who die shortly after hospitalization would not be captured [9]. This may limit the ability to fully capture associations between acute illness severity and Long COVID. Moreover, hospitalized individuals are typically more likely to receive antiviral treatments [22,23], which may reduce the risk of Long COVID, further attenuating any potential state-level associations.

To our knowledge, this study is the first to systematically investigate associations between state-level COVID-19 metrics, health indicators, and adult Long COVID prevalence. We used the best available data sources and aligned the timeframes of these factors with the lag defined in the BRFSS Long COVID survey questionnaire (i.e., symptoms lasting 3 or more months following COVID-19). In addition, this study accounts for variability in the outcome by incorporating the uncertainty inherent in BRFSS survey estimates.

The findings should be interpreted in the context of the study design, data sources and data analyses. First, as an ecological study, we cannot infer causal relationships between state-level factors and Long COVID prevalence. Second, all examined factors except multimorbidity were captured as weekly or monthly aggregates, whereas BRFSS reflects cross-sectional reports of symptoms persisting for 3 or more months. Although we defined the three-month period preceding Long COVID to capture lags, this may still not be adequate to capture the precise lag in individual response. However, we could not use monthly or quarterly survey responses for BRFSS data due to insufficient sample sizes. Further, because BRFSS did not capture information about symptom duration, we were unable to determine if state-level Long COVID burden was driven by newly acquired or ongoing cases (e.g., Long COVID acquired in 2020). Third, differences in testing, reporting practice, access to testing and potential under ascertainment [24] across states may have introduced extra variation in the data sources.

State-level variation in vaccine coverage and cumulative incidence was associated with variation in Long COVID prevalence at the state-level in this ecological study. An emphasis on prevention strategies [25,26] to reduce COVID-19 incidence as well as increasing COVID-19 vaccination coverage may help state and local health departments mitigate the burden of Long COVID in their jurisdictions.

## Data Availability

All relevant data sources are within the manuscript and its Supporting Information files, except for chronic disease prevalence data, which can be obtained upon request.

https://covid.cdc.gov/covid-data-tracker/#datatracker-home

https://www.cdc.gov/nhsn/index.html

https://www.cdc.gov/nchs/hus/sources-definitions/nis.htm

https://www.cdc.gov/brfss/index.html

## Acknowledgements

We thank Dr. Gordana Derado (CDC) and Dr. Diba Khan (CDC) for suggesting the data sources used in our analysis, and Dr. Phillip Salvatore (CDC) and Dr. Ismael Ortega-Sanchez (CDC) for providing valuable input on our analysis.

